# Development and Preliminary Evaluation of a Brief Health Chatbot Acceptability Scale (CAS): A Pilot Study

**DOI:** 10.64898/2026.11.05.26367380

**Authors:** Raymond L. Ownby, Joshua Caballero, Kamilah Thomas-Purcell

## Abstract

**Background:** Conversational agents (chatbots) are increasingly used to deliver health information to middle-aged and older adults, a population in which the determinants of technology acceptance are known to diverge from those of younger users. However, acceptability is inconsistently defined and typically measured with ad hoc, unvalidated instruments. The psychometrically sound chatbot measures that exist assess usability rather than acceptability or were validated in student samples.

**Objective:** To develop and conduct an initial psychometric evaluation of a brief, 10-item health chatbot acceptance measure grounded in the Technology Acceptance Model (TAM) and the Theory of Planned Behavior (TPB).

**Methods:** Twenty adults (mean age = 52.8 years, range 28–81; 85% aged 40 or older; 75% female) enrolled in a sleep-application pilot usability study completed the 10 candidate items, the System Usability Scale (SUS), and sleep measures (PROMIS Sleep Disturbance 8a; Pittsburgh Sleep Quality Index). We examined item performance, internal consistency (Cronbach’s alpha), dimensionality (exploratory factor analysis with oblique rotation), and convergent validity (correlations with the SUS).

**Results:** A two-factor structure emerged and was invariant across rotation methods, resulting in an Information Quality factor and an Acceptance factor (interfactor r = .39). All items loaded ≥ .60 on their primary factor; mean communality was .83. Subscale reliabilities were high (Information Quality α = .96; Acceptance α = .94; full scale α = .93). Both subscales correlated with the SUS in the expected direction (r = .69 and r = .59), providing preliminary support for convergent validity.

**Conclusions:** The measure shows a coherent and theory-consistent two-factor structure and shows preliminary evidence of reliability and convergent validity. Given the small pilot sample, these results must be viewed as provisional but identify the measure as a candidate for formal validation in a larger, independent sample.

## Introduction

### Background

Conversational agents, commonly called chatbots, have moved rapidly from novelty to routine components of digital health delivery. A scoping review of the health care literature identified 47 reports describing conversational agents used for various roles in health care (e.g., treatment and monitoring, health service support, patient education) concluded with a call for rigorous evaluation of their acceptability, safety, and effectiveness (Tudor Car et al., 2020). Other systematic reviews in specific domains have reached similar conclusions: chatbot interventions for physical activity, diet, and weight management show promise but rest on a small and methodologically uneven evidence base (Oh et al., 2021), and reviews of conversational agents for chronic and mental health conditions describe a field still in an early, prototype-dominated stage (Berube et al., 2021; Schachner et al., 2020). Interest in these tools is driven by their scalability, their low threshold for access, and their capacity to deliver behavior-change support outside of scheduled clinical contact.

Middle-aged and older adults are a target population for this work, both because chronic disease burden concentrates in later life and because brain health and dementia risk reduction are increasingly framed as lifestyle interventions requiring sustained engagement. However, the determinants of technology acceptance in this group may differ from those in younger people. The Almere model, developed specifically to explain older adults’ acceptance of assistive social agents, showed that variables relating to social interaction contributed explanatory power beyond perceived usefulness and perceived ease of use (Heerink et al., 2010). A meta-analysis of 35 studies confirmed that perceived usefulness, perceived ease of use, and social influence predict behavioral intention among older adults, while also showing that individuals’ acceptance of unfamiliar health technologies may depend on different factors from more familiar consumer devices (Ma et al., 2021). More recent chatbot-specific work has extended these models further, adding technology anxiety, self-efficacy, perceived physical condition, and self-actualization needs (Wu et al., 2025; Yu & Chen, 2024). In addition, one study of community-dwelling older adults found that stronger anthropomorphism of a social chatbot predicted lower intention to use it (Sou et al., 2026). Taken together, this literature establishes that the determinants of technology acceptance in younger or general-population samples cannot be assumed to be the same in older populations.

Overall, it appears that the term *acceptability* lacks a rigorous definition. In an overview of 43 systematic reviews of health care interventions, Sekhon and colleagues found that none explicitly defined or theorized acceptability, and that most studies inferred it indirectly from behavior such as consent rates, adherence, or dropout (Sekhon et al., 2017). The authors subsequently characterized acceptability as ill-defined, under-theorized, and poorly assessed, and proposed the Theoretical Framework of Acceptability as a multi-construct alternative (Sekhon et al., 2018), later operationalized as a generic questionnaire (Sekhon et al., 2022). A parallel review of the mobile health literature reported that terms such as acceptability, acceptance, and adoption are used inconsistently. Additionally, the authors noted that existing models are rarely adapted to populations with particular needs, and that custom, unvalidated surveys are the predominant measurement approach (Nadal et al., 2020).

These problems are especially acute in chatbot research. In a scoping review of 32 studies evaluating conversational agents for mental health, 203 distinct outcome measurement instruments were identified (Jabir et al., 2023). Nearly three quarters of these instruments appeared in only a single study, and no validity evidence was cited for more than half. Of those, the large majority had been created or adapted specifically for the study in which they appeared. Independent reviews of usability assessment in health conversational agents (Denecke & May, 2022), of voice-based agents for chronic and mental health conditions (Berube et al., 2021), and of chatbots delivering cancer genetic risk assessment (Scalia et al., 2026) have each reported the same pattern of non-comparable, ad hoc measurement. The absence of shared metrics was sufficiently pressing that a Delphi study was convened specifically to build consensus on an evaluation set for health chatbots (Denecke et al., 2021).

Instruments with established psychometric properties exist, but they do not address this issue. The Chatbot Usability Scale (Borsci et al., 2022; Borsci et al., 2023) and the Chatbot Usability Questionnaire (Holmes et al., 2019) are chatbot-specific and psychometrically evaluated; however, both measure usability and satisfaction with interaction rather than acceptability, and both were developed outside of health care contexts. Also, the Artificial Social Agent Questionnaire (Fitrianie et al., 2025) similarly targets interaction experience. Several chatbot acceptance scales have been published recently, but they were developed and validated in higher-education samples of students and teachers (Ibrahim & Nemt-allah, 2026; Polat & Renner, 2026). On the health side, the Artificial Intelligence in Mental Health Scale is brief and psychometrically sound but measures general attitudes toward AI chatbots as a category of mental health support rather than acceptability of a specific deployed intervention (Katsiroumpa et al., 2025), and the Digital Health Acceptability Questionnaire (Haydon et al., 2023), though grounded in the Theoretical Framework of Acceptability (Sekhon et al., 2017), was developed and validated for telephone and videoconference telehealth. The most widely cited study of health chatbot acceptability used a purpose-built survey that does not have readily available psychometric data (Nadarzynski et al., 2019).

In summary, the result is a specific and consequential gap. No brief, theory-grounded, psychometrically evaluated measure is available for assessing the acceptability of health-focused conversational agents among middle-aged and older adults. This is important since this is the population in which acceptance determinants are known to diverge and the population targeted by a growing number of chatbot-delivered health interventions. Investigators developing such interventions are left choosing between long usability instruments that measure something adjacent, acceptance scales validated in students, and locally written items with unknown properties.

### This Study

We report the preliminary psychometric evaluation of a brief measure of chatbot acceptability developed and administered within a development study of Smart Learning and Engagement for Effective Personalized Sleep Using Artificial Intelligence (SLEEP-AI), a behavioral intervention incorporating a conversational agent that delivers tailored sleep-health information to people with sleep difficulties. The measure comprises 10 items drawn from constructs central to the Technology Acceptance Model (TAM) (Davis, 1989) and the Theory of Planned Behavior (TPB) (Ajzen, 1991), selected for brevity so that it could be embedded in a development protocol without adding excessive participant burden. We drew items from TAM and TPB rather than from the Theoretical Framework of Acceptability (Sekhon et al., 2017) because the target construct was intention to use a specific conversational agent rather than the acceptability of a broader care process. The aims of these analyses were to (1) describe item-level performance and distributional characteristics of the 10-item measure in a development sample of 20 participants; (2) estimate internal consistency; (3) examine the measure’s latent structure using exploratory factor analysis; and (4) provide an initial test of convergent validity against an established usability instrument. Consistent with the developmental stage of both the intervention and the instrument, these analyses are preliminary. The sample was determined by the requirements of the development study rather than by a target for scale validation. The exploratory results reported are intended to establish whether the measure performs well enough to warrant formal validation in a larger, independent sample, not to establish the measure as a validated tool.

## Method

### Participants and Procedures

Participants were 20 adults enrolled in a pilot evaluation of a sleep-focused digital application that incorporated an information-delivery chatbot. After completing a usability assessment protocol that included interactions with the chatbot, participants completed the candidate acceptance items along with usability and sleep measures. The sample had a mean age of 52.8 years (SD = 15.5, range 28–81) and was predominantly female (75%). Seventeen participants (85%) were aged 40 years or older, and nine (45%) were aged 50 years or older. Although the measure is intended for middle-aged and older adults, enrollment in the pilot study included persons aged 18 years and older, and three participants were younger than 40 years (aged 28, 33, and 38). Half of participants identified as White (50%), 30% as African-American, and the remainder as Asian, Mixed, or Other. Overall, 35% identified as Hispanic or Latino/a. Consistent with the target application, participants reported clinically meaningful sleep complaints (mean PSQI Global score = 9.1, above the conventional cutoff score of 5 for the presence of sleep difficulties; PROMIS Sleep Disturbance 8a mean T = 54.0), indicating a construct-relevant sample.

Study phases. The pilot was conducted in three sequential phases, with the application and its chatbot revised between phases on the basis of participant feedback and usage, consistent with an iterative development design. Phase 1 (n = 7; June–July 2024) used the initial version of the application and chatbot. The acceptance items were not part of the original protocol. During Phase 1 it became clear that the usability measure did not capture how participants viewed the chatbot as a source of health information, and the 10 candidate items were therefore added to the assessment battery beginning with Phase 2. All participants analyzed here (N = 20) were enrolled in Phase 2 (n = 9; September–December 2024) or Phase 3 (n = 11; August–October 2025); Phase 1 participants completed the SUS but not the acceptance items. The intervention used in Phase 2 was a commercially-available educational website; it was abandoned because of the low usability ratings given by phase 2 participants. The intervention used in Phase 3 included the same verbal content used in Phases 1 and 2 but was incorporated into a purpose-built WordPress website. Phase 2 and Phase 3 participants did not differ significantly in age (M = 55.2 vs. 50.8 years, p = .54). Because the intervention that participants rated was not identical across phases, the pooled sample contains variation attributable to the intervention itself as well as to individual differences. We examined this variation directly by comparing acceptance and usability ratings between phases (see Analytic Approach).

### Measures

#### Health Chatbot Acceptance items

Ten candidate items were developed to represent perceived information quality and credibility (drawing on TAM’s perceived usefulness) and acceptance constructs from the TPB (attitude, subjective norm, perceived behavioral control, and behavioral intention). Each item was rated on a 5-point Likert-type scale (1 = strongly disagree to 5 = strongly agree) with higher scores indicating more favorable perceptions. All items were positively keyed. The full item set appears in the Appendix.

#### System Usability Scale (SUS)

The SUS is a widely used 10-item usability instrument scored 0–100, with higher scores indicating better perceived usability (Brooke, 2013). Standard scoring, including reversal of negatively worded items, was applied. The SUS served as the external anchor for convergent validity for which scores near or above the established average of approximately 68 indicate good perceived usability (Bangor et al., 2008).

#### Sleep measures

The PROMIS Sleep Disturbance 8a short form (Buysse et al., 2010) (reported as a standardized T-score, population M = 50, SD = 10) and the Pittsburgh Sleep Quality Index (PSQI) (Buysse et al., 1989) Global score (0–21; higher scores indicate poorer sleep) characterized the sample and confirmed construct relevance.

### Analytic Approach

#### Descriptive statistics and item analysis

All analyses were completed in SPSS version 29 (Armonk, NY: IBM, Inc.). Item and scale distributions were summarized with means, standard deviations, and observed ranges. For each item we also examined skewness and the proportion of responses at the floor (1) and ceiling (5) of the response scale to identify floor or ceiling effects. Subscale scores were computed as the mean of their constituent items, preserving the 1–5 response metric; the total score was computed as the sum of all 10 items (possible range 10–50). The SUS was scored using standard procedures (Brooke, 2013). Item discrimination was indexed by corrected item–total correlations (the correlation of each item with the sum of the remaining items on its subscale), with values of .30 or greater conventionally considered adequate.

#### Reliability

Internal consistency was estimated with Cronbach’s alpha for the total scale and for each subscale, with 95% confidence intervals computed by Feldt’s method (Feldt et al., 1987), along with alpha-if-item-deleted to identify items whose removal would raise reliability. Alpha values of .70 or greater were considered adequate for research use (Boateng et al., 2018). Because alpha increases with inter-item correlation, values above approximately 0.95 were interpreted as possible item redundancy rather than as evidence of superior measurement.

***Dimensionality*** was examined with exploratory factor analysis using principal axis factoring. Factorability was evaluated with the Kaiser-Meyer-Olkin (KMO) measure of sampling adequacy and Bartlett’s test of sphericity. Both orthogonal (varimax) and oblique (direct oblimin) rotations were examined. Since the theory does not require the factors to be uncorrelated, the oblique solution is reported as primary and the interfactor correlation is presented. Individual response patterns were inspected for atypical cases, whose influence on the solution was examined by repeating the analysis with them excluded.

***Convergent validity*** was assessed with Pearson correlations between the full scale and derived subscale scores (computed as item means) and the SUS total.

#### Phase comparisons

Because the intervention was revised between Phases 2 and 3, total and subscale acceptance scores and SUS totals were compared between phases using independent-samples *t* tests with Welch’s correction for unequal variances. Given the small group sizes and the negatively skewed distributions, Mann– Whitney U tests were run as a sensitivity check. Effect sizes are reported as Hedges’ g (positive values indicate higher scores in Phase 3). SUS scores, which were also collected in Phase 1, were compared across all three phases with one-way ANOVA and the Kruskal-Wallis test. To determine whether phase differences in acceptance and usability moved together, and whether pooling across phases inflated the convergent validity estimates, we compared the acceptance–SUS correlations within each phase with the pooled correlations and computed partial correlations between acceptance and SUS controlling for phase.

#### Interpretability of a small-sample factor analysis

The adequacy of a sample for factor analysis depends less on the raw case-to-item ratio than on the level of communality and the degree to which factors are overdetermined (i.e., defined by several strongly loading items). Under conditions of high communality and well-overdetermined factors, factor solutions recover population structure well even at small sample sizes (MacCallum et al., 1999), and a factor defined by four or more loadings above .60 may be stable largely independent of sample size (Guadagnoli & Velicer, 1988). These conditions are used below to assess the interpretability of the present solution.

## Results

### Descriptive Statistics

Table 1 presents descriptive statistics for the sample characteristics, the derived acceptance subscales, and the external measures. Both acceptance subscales had means well above the scale midpoint, and the SUS mean (78.6) exceeded the commonly cited average usability benchmark of 68 (Bangor et al., 2008), consistent with a favorably received application. The wide SUS standard deviation (25.4; range 10– 100) likely reflects the pooling of values across two study phases that assessed different versions of the intervention.

**Table 1.** Descriptive Statistics (N = 20)

| **Measure** | **Mean** | **SD** | **Minimum** | **Maximum** |
|---|---|---|---|---|
| Age (years) | 52.80 | 15.50 | 28 | 81 |
| PROMIS Sleep Disturbance 8a (T-score) | 54.02 | 2.53 | 46.7 | 58.1 |
| PSQI Global (0–21) | 9.10 | 2.69 | 5 | 13 |
| SUS total (0–100) | 78.63 | 25.37 | 10 | 100 |
| Chatbot total (sum, 10 items) | 40.75 | 8.03 | 23 | 50 |
| Information Quality subscale (mean) | 4.22 | 0.94 | 1.20 | 5.00 |
| Acceptance subscale (mean) | 3.93 | 0.93 | 1.20 | 5.00 |

*Note.* Subscale scores are item means on the 1–5 response metric. PSQI = Pittsburgh Sleep Quality Index; PROMIS = Patient-Reported Outcomes Measurement Information System; SUS = System Usability Scale.

At the item level, the responses of all 10 items were negatively skewed (skewness −0.45 to −2.26), with 25% to 55% of responses at the scale ceiling and no more than one response (5%) at the floor for any item. Ceiling effects were most pronounced for the understanding, belief, and trust items (50–55% at ceiling) and least pronounced for the subjective-norm item (25%).

### Reliability

Internal consistency was high for the total scale (Cronbach’s α = .93, 10 items) and for both subscales (Information Quality α = .96, 5 items; Acceptance α = .94, 5 items). The very high alpha for the Information Quality subscale likely reflects redundancy among its items as the belief and trust items were correlated at *r* = 0.98, as alpha increases with greater inter-item correlations. The 95% confidence intervals were 0.87-0.97 for the total scale, 0.92-0.98 for Information Quality, and 0.89-0.97 for Acceptance. Corrected item–total correlations ranged from 0.82 to 0.93 within the Information Quality subscale and from 0.75 to 0.91 within the Acceptance subscale. Only two deletions would have raised a subscale’s alpha. If the relevance item were deleted from Information Quality α would change from 0.956 to 0.961 and if the subjective-norms item were deleted from Acceptance α would change from 0.942 to 0.946.

### Factorability and Factor Structure

The correlation matrix was suitable for factor analysis with a KMO = .705, above the conventional .60 threshold, and Bartlett’s test of sphericity was significant, χ²(45) = 249.01, p < .001. Principal axis factoring yielded two factors. The solution was invariant to rotation (varimax vs. oblimin), consistent with a stable structure.

Inspection of individual response patterns identified one participant who disagreed or strongly disagreed with all five Information Quality items while agreeing or strongly agreeing with four of the five Acceptance items. With this case excluded (n = 19), the correlation between the subscales rose from *r* = .47 to *r* = .80, and the Information Quality items continued to form a separate factor, although the second factor was considerably weaker; internal consistency was essentially unchanged (Information Quality α = .91; Acceptance α = .95).

The oblique (direct oblimin) pattern matrix is presented in Table 2. Factor 1, Information Quality, was defined by items concerning comprehension, relevance, belief, trust, and the perceived usefulness of the information for improving health. Factor 2, Acceptance, was defined by items concerning enjoyment, positive attitude, social approval, confidence in use despite problems, and intention to reuse the chatbot. All items loaded ≥ .60 on their intended factor, and cross-loadings under the oblique rotation were minimal. The two factors were moderately correlated (r = .39), indicating related but separable dimensions and justifying the oblique rotation. Additionally, an orthogonal rotation of the same data reproduced the identical item-to-factor assignment.

**Table 2.** Pattern Matrix (Principal Axis Factoring, Oblimin Rotation) and Communalities.

| Item | F1: Info. Quality | F2: Acceptance | *h²* |
|---|---|---|---|
| Understood the information | .99 | −.12 | .90 |
| Information was relevant/what I asked | .73 | .28 | .76 |
| Believe the information | .98 | −.08 | .91 |
| Trust the information | .97 | −.06 | .89 |
| Could use information to improve health | .79 | .26 | .85 |
| Enjoyed interacting with the chatbot | .09 | .85 | .78 |
| Positive attitude toward the chatbot | .03 | .92 | .87 |
| Friends/family would approve | .35 | .65 | .71 |
| Could use chatbot despite problems | −.12 | .90 | .75 |
| Would use chatbot again | −.06 | .98 | .91 |

*Note.* Primary loadings in each row are the larger of the two. *h²* = extraction communality. Mean *h²* = .83; the two factors together accounted for approximately 83% of total item variance. Interfactor correlation *r* = .39.

### Convergent Validity

Both subscales were correlated with the SUS in the predicted direction and at moderate-to-strong sizes: Information Quality, *r* = .69, p < 0.001; Acceptance, *r* = .59, *p* = 0.006 (N = 20). These correlations are large enough to demonstrate that the acceptance measure tracks independently assessed usability. The two subscales were correlated with each other at *r* = .47 (*p* = 0.035), closely matching the interfactor correlation from the factor analysis (*r* = .39).

#### Phase Comparisons

Acceptance and usability scores by phase are presented in Table 3. Total acceptance scores were significantly higher in Phase 3 (M = 44.45, SD = 6.31) than in Phase 2 (M = 36.22, SD = 7.84), *t*(15.3) = 2.55, p = 0.022, *g* = 1.12. Both subscales showed differences of similar size (Information Quality *g* = 0.88; Acceptance *g* = 0.95). These reached significance on Mann–Whitney tests (*p* = .040 and *p* = .035) but were not significant on Welch’s *t* test (*p* = .076 and *p* = .053). At the item level, the largest differences were for relevance of the information (*g* = 1.26), subjective norm (*g* = 1.31), and understanding (*g* = 1.04). Belief, trust, and intention to reuse differed least (*g* = 0.42–0.53). Phase 3 scores were also more homogeneous: the standard deviation of the total score fell from 7.84 to 6.31 and that of the Information Quality subscale from 1.11 to 0.62.

**Table 3.** Acceptance and Usability Scores by Study Phase.

| **Measure** | **Phase 2** | **Phase 3** | ***t* (df)** | ***p*** | ***g*** |
|---|---|---|---|---|---|
| Acceptance total (sum, 10 items) | 36.22 (7.84)<sup>1</sup> | 44.45 (6.31)<sup>1</sup> | 2.55 (15.3) | 0.022 | 1.12 |
| Information Quality (mean) | 3.78 (1.11) | 4.58 (0.62) | 1.94 (12.0) | 0.076 | 0.88 |
| Acceptance (mean) | 3.47 (1.02) | 4.31 (0.67) | 2.12 (13.3) | 0.053 | 0.95 |
| SUS total (0–100) | 63.89 (30.90) | 90.68 (10.19) | 2.49 (9.4) | 0.033 | 1.17 |

<sup>1</sup> Values are means and standard deviations

Usability showed a parallel pattern. SUS scores were essentially unchanged from Phase 1 (M = 63.93, SD = 19.73, n = 7) to Phase 2 (M = 63.89, SD = 30.90) but rose sharply in Phase 3 (M = 90.68, SD = 10.19), Phase 2 vs. Phase 3 t(9.4) = 2.49, p = .033, g = 1.17; across all three phases, F(2, 24) = 5.09, p = .014 (Kruskal–Wallis H = 10.77, p = .005). The Phase 3 mean is well above the commonly cited benchmark of 68 (Bangor et al., 2008), whereas the Phase 1 and Phase 2 means fell slightly below it.

Because acceptance and usability both increased in Phase 3, part of their pooled association could reflect the differences of scores across phases. Controlling for phase reduced, but did not eliminate, the convergent correlations: partial r = .60 (p = .006) for Information Quality and .45 (p = .051) for Acceptance, compared with pooled r = .69 and .59; for the total score, the correlation fell from r = .75 to partial r = .65 (p = .003). Within phases, correlations between the total score and the SUS were r = .81 (p = .008) in Phase 2 and r = .36 (p = .27) in Phase 3. The weaker Phase 3 association is consistent with the restricted range of both measures in that phase, with most participants near the ceiling on each.

## Discussion

This pilot study developed a brief 10-item measure of health chatbot acceptance grounded in the TAM and TPB frameworks and examined its psychometric properties in a small sample of predominantly middle-aged and older adults. The measure showed a coherent two-factor structure (i.e., Information Quality, Acceptance) that was invariant across rotation methods, provided high internal consistency, and produced convergent validity against an established usability instrument. The moderate correlation between the two factors indicates that perceptions of information quality and behavioral acceptance are related but separable, consistent with the theoretical distinction between the credibility and usefulness of delivered information and the disposition to adopt and continue using the agent.

The observed structure is theoretically interpretable. Factor 1 aligns with the TAM’s dimension of perceived usefulness, extended to include the comprehension, credibility, and trustworthiness of health information. These dimensions are especially salient when an automated agent delivers health content, and when users are older adults for whom trust and comprehension are recurrent themes in the acceptance literature (Heerink et al., 2010; Ma et al., 2021; Sou et al., 2026). Factor 2 collapses several TPB constructs (i.e., attitude, subjective norm, perceived behavioral control, behavioral intention) into a single acceptance dimension. That these TPB constructs did not separate empirically is unsurprising given the use of a single indicator per construct. The subjective-norm item (“friends/family would approve”) was the most complex, showing the largest secondary loading, consistent with subjective norm being the TPB construct least redundant with attitude and intention. Although we drew items from TAM and TPB, the two emergent factors map broadly onto constructs in the Theoretical Framework of Acceptability (Sekhon et al., 2017). These included *Information Quality* onto perceived effectiveness and intervention coherence, and *Acceptance* onto affective attitude and self-efficacy.

The between-phase comparison helps explain the wide spread of scores in this small sample. Participants in Phases 2 and 3 rated different versions of the study intervention, and both acceptance and usability were substantially higher, and less variable, in Phase 3, with effect sizes above one standard deviation. Part of the variability in the pooled sample therefore reflects changes in the intervention rather than individual differences in disposition toward chatbots. This has two implications. First, it is encouraging for the measure’s intended use in formative evaluation in that the scale appears sensitive to changes in the intervention participants were rating. Further, those changes paralleled independently assessed usability on the SUS. The items that changed most (relevance, understanding, and subjective norm) are plausibly those most affected by changes in the intervention, whereas belief and trust in the information were high in both phases and changed least.

### Limitations

The principal limitation of this exploratory study is the small sample (N = 20). The conditions under which previous reports have shown that analyses using small samples may recover factor structure were met, with high item communalities and factors determined by five items each with loadings greater than 0.60 (Guadagnoli & Velicer, 1988; MacCallum et al., 1999). Given the small sample, however, these results remain preliminary. The very high inter-item correlations produced a near-singular correlation matrix, reflecting redundancy among the most similar items, belief and trust. As a result, this suggests that near-duplicate items should be consolidated in a revised version of the scale. The separation between the two factors was sensitive to a single respondent with an atypical response pattern. With this case excluded, the subscale intercorrelation rose from r = .47 to r = .80. The factor structure should therefore be retested in an independent sample.

Finally, the sample was drawn from a single sleep application pilot study and was demographically specific. Although the measure is intended for middle-aged and older adults, the sample included three participants younger than 40 and eight in their 40s, and was too small to test whether the measure functions equivalently across age groups. Generalizability to other chatbots, populations, and health domains is unknown.

### Future Directions

Confirmatory factor analysis was not appropriate for these data since fitting a two-factor confirmatory model to the same sample that generated the structure would be circular, and the model’s parameter count exceeds the available cases (Wolf et al., 2013). The necessary next step is replication in an independent, adequately powered sample, in which the pre-specified two-factor structure can be tested to evaluate the solution found in this pilot. A revised item set that consolidates redundant Information Quality items and strengthens the underrepresented subjective-norm content with additional items would improve the measure. Following best-practice scale-development guidance (Boateng et al., 2018), subsequent work should add test-retest reliability, discriminant validity, and evaluation of measurement invariance across age. An a priori power analysis for the confirmatory model, for example by Monte Carlo simulation using the present solution as the population model, may be useful in determining the sample size required for that next stage.

## Conclusion

A brief, theory-based measure of health chatbot acceptance suggested a coherent two-factor structure, preliminary reliability, and convergent validity against an established usability scale in a pilot sample predominantly including middle-aged and older adults. These provisional findings support the measure as a candidate instrument for formative evaluation of health chatbots in this population and provide a basis for confirmatory testing in future, adequately powered research.

## Declarations

### Ethics

This study was approved by the Institutional Review Board of Nova Southeastern University (protocol 2024-216, approved April 29, 2024). All participants provided written informed consent.

### Data availability

The full item set and data are available from the first author on reasonable request.

### Use of generative AI

Claude Opus 5.5 (Anthropic) was used to assist with text drafting and editing, supplementary statistical analyses, and reference checking. All analyses were verified by the authors, who reviewed all content and take full responsibility for the manuscript.

### Related work

Usability and qualitative findings from the same pilot are reported in a separate mixed-methods manuscript, in preparation.

## Appendix. Chatbot Acceptance Scale (CAS) Items

Response scale: 1 = strongly disagree, 2 = disagree, 3 = neither agree nor disagree, 4 = agree, 5 = strongly agree. All items are positively keyed; no reverse scoring. Subscale scores are computed as the mean of their five items.

### Factor 1 — Information Quality

1. I understood the information the chatbot gave me about my health.
2. The information the chatbot gave me was exactly about what I had asked it.
3. I believe the information the chatbot gave me.
4. I trust the information the chatbot gave me.
5. I could use the information the chatbot gave me to improve my health.

### Factor 2 — Acceptance

6. I enjoyed interacting with the chatbot.
7. I really liked working with the chatbot.
8. My friends and family would approve of me getting information from the chatbot.
9. I would be able to use the chatbot even if I ran into problems with it.
10. I would use the chatbot again to get health information.

